# Comparison the outcomes of blunt trauma with penetrating traumatic popliteal artery injury repair in Southwest of Iran

**DOI:** 10.1101/2023.06.10.23291220

**Authors:** Alireza Azadvar, Seyed Masood Mousavi, Hossein Minaie Tork, Shaghayegh Sherafatmand, Hossein Ghaedamini

## Abstract

**Introduction:** Popliteal artery injuries (PAI) one of the most severe peripheral vascular damages may lead to movement impairment or amputation. This study aimed to compare the repair results of popliteal artery injury due to blunt trauma with penetrating trauma in patients referred to the Vascular Surgery Center in Southwest Iran.

**Methods:** This was a descriptive and retrospective study. The statistical population was all patients with vascular trauma referred to the Southwest Trauma Center of Iran in 2020. The sampling method was census. The data collection tool was a checklist containing demographic information, medical information, vascular information, Functional Independence Measure (FIM) Score, and Mangled Extremity Severity Score (MESS). Statistical analyses were conducted using SPSS24 (IBM Inc., Chicago, IL, USA). The Chi-Square test, Fisher Exact Test, and independent T-test were used.

**Results:** 63.1 % of participants had penetrating trauma. 10.6 % of participants needed limb fasciotomy (26.3 % in the penetrating trauma group VS 1.7 % in the blunt trauma group). The amputation rate was 8.8 % in the penetrating trauma group VS 12 % in the blunt trauma. There was a significant difference between the two groups in terms of organ fasciotomy (P=0.035), type of PAI(P=0.018), and fracture (P=0.007). MESS Score (P<0.001), FIM Score (P<0.001), time of discharge (P<0.001), and Arterial condition (P<0.001).

**Conclusion:** Popliteal artery trauma is uncommon. Tthe results showed that the complication of penetrating PAI include organ fasciotomy (type of fracture, MESS Score, FIM Score, time of discharge, Arterial condition) in the southwest of Iran were significantly more than the blunt type. It is necessary to investigate additional studies with a larger sample size and extended duration.

## Introduction

Vascular trauma is one of the most dangerous types of injuries that require a quick and reliable diagnosis (1-3). The prevalence of vascular trauma is 8-10 % caused by traffic accidents, gunshots, shotguns, and stab wound injuries (4-6). Immediate surgeries (exploration, anastomosis, ligation, graft) is the best treatment (2).

Among all lower extremity vascular injuries, Popliteal artery injuries (PAI) one of the most severe peripheral vascular damages may lead to movement impairment or amputation (7,8). Acute ischemia is one important side effect of PAI, which can lead to amputation or severe long-term complications (7-10).

We have just less than 6 hours times to prevent ischemia and re-establish blood circulation in the limb to prevent permanent injury, although it depends on other factors such as the level of injury, previous arterial disease, soft tissue trauma, and previous history of lower limb surgery (11,12).

The difference in outcomes of blunt and penetrating popliteal artery trauma is an issue that has always been of investigating by vascular surgeries (11-14). The results of Banion et.al showed that there is no difference between the outcome of popliteal artery injury due to blunt and penetrating trauma (15). The results of Zhong et.al showed that technical success rate of endovascular repair of blunt popliteal arterial injuries was 100% (16).

Due to limited studies all over the world about the difference in the outcomes of blunt and penetrating popliteal artery trauma and that no study has been conducted in southwest Iran, the present study aims to compare the outcomes of blunt trauma with penetrating traumatic popliteal artery injury repair in Southwest of Iran that we can prevent the possible complications of each type of PAI.

## Methods

This was a descriptive and retrospective study. The statistical population was all patients with vascular trauma (penetrating and Blunt) referred to the Southwest Trauma Center (Golestan Hospital in Ahwaz related to Joundishapur university of medical sciences, a large Level I urban trauma center) during 2020. Sampling method was census.

The Inclusion criteria was popliteal artery injuries and need for emergency surgery and the presence of it was confirmed in Operating Room (OR). The exclusion criteria were Incomplete documents and the death of patients before surgery.

The data collection tool was a checklist containing demographic information (age, gender), Systolic Blood Pressure (SBP), Diastolic Blood Pressure (DBP), Glasgow Coma Scale (GCS), Comorbidities (Diabetes, Hypertension, Coronary artery disease, Active Smoker), preoperative antiplatelet therapy (Aspirin, Plavix), organ Fasciotomy, type of popliteal artery injury, type of fracture, concomitant soft tissue damage, concomitant nerve damage, concomitant venous injury, need for angiography, Ischemic time, time of hospitalization, time of ICU admission, time of surgery, Functional Independence Measure (FIM) Score, Mangled Extremity Severity Score (MESS), patient status at the time of discharge, arterial condition, need for amputation.

The long-term consequences of surgery are measured with FIM and the degree of disability, and the factors affecting it. The FIM is a disability assessment scale for post-discharge victims that is divided from grade 4 (complete dependence on companions) to grade 1 (independent) and in cases where it is greater than or equal to 6, the degree of severe functional disability is considered. At point 1, the patient is able to walk 150 meters independently of others without assistive devices (mild disability). In score 2, the patient is able to walk independently of others but with aid of 150 meters or more (moderate disability), in score 3, the patient is dependent on the help of others but is able to walk with the help of others and with aids of 50 meters (Moderate disability). In score 4, the patient is dependent on the help of others, and cannot walk even 50 meters with assistive devices (severe disability). Based on the score, it is decided whether to amputate or keep the limb, which is a recommendation for amputation at a score above 7 (17-19).

The severity of the vascular injury was measured by the MESS scoring system that combined vascular and orthopedic injuries and used to select lower-extremity injuries that warrant a primary amputation. Vascular injury was never clearly defined in the MESS scoring system, and the MESS score allows for the evaluation of patients with normal perfusion. For this reason, the MESS has been widely referenced as the trauma limb-salvage index for lower-extremity trauma. Four characteristics related to injury: Injury severity (energy transfer), Ischemia (20).

Quantitative and categorical data were expressed as mean (SD) (median, minimum and maximum) and frequency (percentage), respectively. Statistical analyses were conducted using SPSS_24_ (IBM Inc., Chicago, IL, USA). The Chi-Square test, Fisher Exact Test abd independent T test used for categorical and quantitative data. The research protocol was approved by the Medical Ethics Committee of Joundishapor University of Medical Sciences (R.AJUMS.HGOLESTAN.REC.1399.087).

## Results

92 participants were included in the study, of which 58 (63.1 %) had penetrating trauma. There is no difference between two groups according to age (P>0.05), systolic blood pressure (SBP) (P>0.05), diastolic blood pressure (DBP) (P>0.05), Glasgow Coma Scale (GCS) (P>0.05), Comorbidity (P>0.05) and preoperative antiplatelet therapy (P>0.05) (Table 1).

**Table 1.** Basic characteristics of Participants

| Variable |  | Blunt Trauma<br>(N=58) | Penetrating<br>Trauma<br>(N=34) | P.V |
| --- | --- | --- | --- | --- |
| | | Mean ± S.D | Mean ± S.D | |
| Age | | 45.37 ± 9.14<br>(19-80) | 49.43 ± 10.65<br>(18-77) | 0.614 |
| SBP | | 11.63 ± 2.9 | 11.1 ± 3.44 | 0.361 |
| DBP | | 9.34 ± 3.7 | 9.5 ± 0.9 | 0.840 |
| GCS | | 10.8 ± 1.3 | 11.72 ± 0.7 | 0.214 |
| Male Gender |  | 64.7 (22) | 67.2 (39) | 0.540 |
| Comorbidity | Diabetes | 11.7(4) | 5.1(3) | 0.251 |
|  | Hypertension | 20.5(7) | 13.7(8) | 0.459 |
|  | Coronary artery<br>disease | 2.9(1) | 1.7(1) | - |
|  | Active Smoker | 41.1 (14) | 34.4(20) | 0.513 |
| Preoperative<br>antiplatelet<br>therapy | Aspirin | 17.6 (6) | 18.9 (11) | 0.415 |
|  | Plavix | 5.8 (2) | 6.8 (4) | 0.187 |
|  | None | 76.6 (26) | 74.3 (43) | 0.318 |

SBP: Systolic Blood Pressure
DBP: Diastolic Blood Pressure
GCS: Glasgow Coma Scale

10.6 % of participants needed limb fasciotomy (26.3 % in penetrating trauma group VS 1.7 % with blunt trauma group). The amputation rate was 8.8 % in penetrating trauma group VS 12 % in blunt trauma. There was a significant difference between two group in the term of organ fasciotomy (P=0.035), type of PAI(P=0.018), fracture (P=0.007). MESS Score (P<0.001), FIM Score (P<0.001), time of discharge (P<0.001), Arterial condition (P<0.001), (Table 2).

**Table 2.** Consequences of trauma injury of Participants

| Variable |  | Penetrating Trauma<br>(N=34) | Blunt Trauma<br>(N=58) | P.V |
| --- | --- | --- | --- | --- |
|  |  | N (%) | N (%) |  |
| Organ Fasciotomy | No | 25 (73.7) | 57 (98.3) | 0.035 |
|  | Primary | 2 (5.8) | 0 (0) |  |
|  | Therapeutic | 7 (20.5) | 1 (1.7) |  |
| Type of popliteal artery injury | Proximal | 18 (53) | 37 (63.9) | 0.018 |
|  | Middle | 9 (24.6) | 3 (5.2) |  |
|  | Distal | 7 (13.2) | 18 (31) |  |
| Fracture | Hip | 0 (0) | 2 (3.4) | 0.007 |
|  | Tight | 1 (2.9) | 5 (8.6) |  |
|  | Knee | 0 (0) | 12 (20.6) |  |
|  | Calf | 3 (8.8) | 27 (46.5) |  |
| Concomitant Soft Tissue Damage |  | 11 (32.3) | 4 (6.9) | 0.798 |
| Concomitant Nerve Damage |  | 3 (8.8) | 1 (1.7) | 0.753 |
| Concomitant Venous Injury |  | 4 (11.7) | 3 (5.2) | 0.97 |
| Need for angiography |  | 3 (8.8) | 5 (8.6) | 0.064 |
| - | | Mean ± S.D | Mean ± S.D | - |
| Ischemic Time (Hours) | | 5.7 ± 1.4 | 6.3 ± 2.6 | 0.91 |
| Time of Hospitalization (day) | | 22.4 ± 2.3 | 24.9 ± 1.6 | 0.64 |
| Time of ICU admission (day) | | 13.8 ± 2.1 | 15.9 ± 1.4 | 0.42 |
| Time of Surgery (Hours) | | 3.2 ± 0.9 | 3.7 ± 0.4 | 0.07 |
| MESS Score | | 6.5 ± 0.7<br>(6-11) | 5.3 ± 0.5<br>(3-9) | <0.0001 |
| FIM Score | 1 | 13 (38.2) | 26 (44.9) | <0.0001 |
|  | 2 | 21 (61.8) | 32 (55.1) |  |
| Patient status at the time of discharge | Complete Recovery | 21 (61.8) | 41 (70.8) | <0.0001 |
|  | Partial Recovery | 10 (29.4) | 15 (25.8) |  |
|  | Expire | 3 (8.8) | 2 (3.4) |  |
| Arterial condition | Intimal flap | 1 (2.9) | 6 (10.3) | <0.0001 |
|  | spasm | 0 (0) | 3 (5.1) |  |
|  | Complete Cut | 7 (20.6) | 14 (24.5) |  |
|  | Thrombosis | 24 (70.6) | 32 (55.1) |  |
|  | Tearing | 2 (5.9) | 3 (5.1) |  |
| Amputation | Minor | 2 (5.9) | 6 (10.3) | 0.45 |
|  | Major | 1 (2.9) | 1 (1.7) |  |

## Discussion

This was the first study that investigated the outcome of PAI in Southwest of Iran. PAI s are uncommon even in busy urban trauma Centers and little data are available in southwest of Iran about functional outcome after it.

PAI are often seen as fractures, dislocations or penetrating injuries (4). Concern for this problems is very important for organ preservation. Also, the time when patients arrive at the hospital can lead to a reduction in injuries. Most blunt popliteal injuries are not easily diagnosed and are dangerous (unlike penetrating injuries.)

The mean score of MESS was statistically higher in penetrating trauma group which is similar to the result of Assieno et al (11) (MESS in blunt trauma was 5.62 and in penetrating trauma was 5.86) and against the result of Simons et al (12) (MESS in blunt trauma was 6.52 and in penetrating trauma was 4.92). The reason for the difference in the results can be due to the verity in sample size, the statistical population, inclusion and exclusion criteria. Also, this research was conducted in southeast of Iran where the rate of accidents, crimes and fights is very high (22).

The study of Simmons et al (12) showed that a MESS more than 7 is the best independent prediction of amputation; however, 63 % of patients with MESS more than 7 did not require an amputation. This suggests that MESS is predictive of amputation but should not be the only factor that determines the need for amputation after a popliteal artery injury. But this issue was not investigated in the present study. We suggest that future studies in southwest of Iran evaluate the predictivity of MESS for amputation.

The mean score of FIM was also statistically higher penetrating trauma group which is similar with the study of Nikolaus et al and against with the study of Mulinex and et al (14). the difference may be related to the limitations of the FIM score (nontrauma-specific design and the effect of pre-existing disabilities in the assessment).

There wasn’t any significant difference in amputation rate between the two groups that is similar with but Dua et al identified that blunt injury combined with fractures and an ISS (Injury Severity Score) > 9 is associated the risk of amputation, whereas in a military group, Penetrating injuries due to explosive devices are associated with worse functional outcome and a higher amputation (24). Obniaon et al (15) showed that amputation risk is higher in blunt popliteal injury compromised with penetrating group which is similar with results of Sciarretta et al. The results showed that multiple risk factors for amputation include MESS score <7, long ischemic time, pulseless distal limb, associated venous injury, severed nerves, open fractures, blunt injury, shock, and advanced age.

The results showed that there was no significant association between patients’ BMI and amputation rate, which was inconsistent with the study of Simmons et al. They showed that BMI in the blunt trauma group is significantly higher than the penetrating group. The prevalence of obesity is increasing in Iran. Obesity is an independent risk factor in various diseases, especially trauma. The results of Simons et al. showed that a BMI more than 40 can lead to lower limb amputation (23).

The result shows there isn’t any difference in fasciotomy rate between two groups that similar to the result of Kauvar et al (25), Mulinex et al (14) and Sciarretta et al (26). It can be concluded that performing fasciotomy don’t have any association with the type of trauma.

The results showed that there isn’t any difference in time of Hospitalization (day) and time of ICU admission (day) between two groups but the study of Sciarretta et al (26) showed the time of Hospitalization and ICU admission are longer in blunt group.

The result showed there isn’t any difference in Ischemic Time rate between two groups bout many studies showed that maintaining ischemic times as close to six hours may lead to improved outcomes for these difficult and rare injuries (11-14).

The result showed that Patient status at the time of discharge is better in blunt PAI that is not similar to the result of Cooper et al (27), Sciarretta et al (26), Mulinex et al (14), Kauvar et al (25). They verified that blunt trauma has a significantly worse outcome if compared with penetrating trauma. Complementary research should confirmed about this subject in southeast of Iran. It may be due to the fact that in the present study, the patients did not only have problems with the popliteal vessels (pure vascular problems) and had more trauma on other parts of their body as well (such as the brain, chest, and abdomen), which affects their prognosis.

The results showed that the rate of fasciotomy in blunt trauma was higher than in penetrating trauma, which was similar with the results of Sciarretta et al (26).

This study has several limitations like small sample size, inhomogeneous in trauma mechanism, different imaging of PAI and variables like ISS, Dislocation, Limb Salvage Index, Initial base deficit, technique of fasciotomy. In this study we couldn’t evaluate the functional status of extremities after PAI.

## Conclusion

Popliteal artery trauma is relatively uncommon. Totally, the results showed that the complication of penetrating PAI include organ fasciotomy, type of fracture (P=0.007). MESS Score, FIM Score, time of discharge, Arterial condition, in the southwest of Iran were significantly more than the blunt type. Our amputation rate was 9.1 % with no significant difference in rates between penetrating and blunt trauma (8.8% vs 12%). It is necessary to investigate additional studies with a larger sample size and extended duration.

## Data Availability

All data produced in the present study are available upon reasonable request to the authors

## Author’s contributions

MM conceived of the present idea; AA developed the idea and written informed consent from the patient, and HG wrote the manuscript. HM helped to conduct the literature review, verified the analytical methods, and wrote the manuscript. HG also helped to conduct the literature review. MM performed the experiments, whereas HG contributed to the analysis and wrote the manuscript. AA helped with the analyses and wrote the manuscript. All authors reviewed the results and approved the final version of the manuscript.

## Acknowledgments

The authors would like to thank the patients and personnel of Golestan hospital for their cooperation in performing the present study.

